# Management of possible drug-drug interactions in medical prescriptions received in pharmacies

**DOI:** 10.1101/2025.06.24.25330202

**Authors:** Carelle D Nnanga, Donald Embogo, AP Minyem Ngombi, Eric Nseme

## Abstract

**Introduction:** Pharmacists and their community pharmacy are responsible for analysing medical prescriptions to identify possible drug-drug interactions. In Cameroon, drug-drug interactions have been examined on several occasions in the hospital settings, but rarely in the community pharmacies, especially from the point of view of management practices. The aim of this study was to describe the management of potential drug-drug interactions in prescriptions received in pharmacies in the Efoulan Health District, Yaoundé, Cameroon.

**Methodology:** A cross-sectional descriptive study with prospective data collection was conducted over a period of 07 months from November 2022 to May 2023 in all pharmacies in the Efoulan Health District. All legible prescriptions containing at least two drugs were included. Data were collected using a pre-established and pre-tested form. Interactions were identified using the THERIAQUE® drug database. Data analysis was performed using IBM-SPSS® Version 23.0 software.

**Results:** The study involved 67 prescriptions containing 124 drug interactions, averaging 1.8 interactions per prescription. The interactions were mainly pharmacodynamic (74.2%) versus pharmacokinetic (25.8%) in terms of type, and to be taken into account in terms of risk level (52.5%). They were mainly formed by synergism for pharmacodynamic interactions (89.1%) and complexation for pharmacokinetic interactions (59.4%). Prescribers were mainly cardiologists (42.9%) and general practitioners (42.9%). These pharmacological reactions occurred mainly between two non-steroidal anti-inflammatory drugs. The detection rate for interactions was low (21.8%) due to the use of personal knowledge (85.2%) and physical documents (14.8%) for detection. The interactions mainly detected were pharmacokinetic (13.7%) and the combinations to be used with caution (13.7%). The attitudes adopted were, in descending order: advise the patient (55.5%), do nothing (40.8%) and refer the patient back to the prescriber (3.7%).

**Conclusion:** Many drug interactions can occur as a result of medical prescriptions. Few of them are detected by dispensers, and the resulting attitudes vary.

## INTRODUCTION

Medical prescription is a clinical process whereby a treatment is written on a document called a “prescription”, which serves as the primary means of communication between the prescriber and the pharmacist [1]. From a regulatory standpoint, a medical prescription is the outcome of an organised process and constitutes a forensic medical document involving both civil and criminal liability for the physician and the pharmacist [2]. Drug interactions can occur when prescribed drugs interact with each other, with food or with nutritional supplements. These are pharmacological, pharmacokinetic or clinical reactions resulting from an alteration (either enhancement or reduction) in the effect of one drug due to the prior or concomitant use of another substance [3]. They can lead to non-compliance, therapeutic failures or potentially serious accidents that can compromise the prognosis, particularly in cases of polymedication [4]. Regulatory guidelines require that, once a prescription is received in a pharmacy, the pharmacist must carry out a pharmaceutical analysis to detect and address any medication-related errors, including drug-drug interactions (DDIs) [5]. It is estimated that 42% of adverse drug events are preventable and occur mainly during prescribing, a large proportion of which are due to DDIs [6]. They can be found in both hospitals and community pharmacies. Numerous studies worldwide have examined drug interactions in hospitals, but little in pharmacies, and even less on the management methods used. In 2021, a study carried out at the Jamot Hospital in Yaoundé, Cameroon, showed that nearly 80% of prescriptions for psychotropic drugs included at least three drug interactions [7]. In light of this important issue, we undertook this study with the objective of describing how potential drug interactions are managed in medical prescriptions received in community pharmacies in the Efoulan Health District, Yaoundé, Cameroon.

## MATERIAL AND METHODS

A cross-sectional descriptive study with prospective data collection was conducted in all community pharmacies in the Efoulan Health District, in Yaoundé, over a period of 7 months, from November 1st 2022 to May 25th 2023. Our study population consisted of all medical prescriptions received in those pharmacies. All prescriptions containing at least two drugs with identification of the patient and prescriber present and having obtained informed consent signed by the patient were included. We excluded all prescriptions that were not legible. Data were collected using a pre-established and pre-tested anonymous form. Prescription interactions were identified using the THÉRIAQUE® drug database. This is a database of all drugs available in France, intended for healthcare professionals. Data were entered and analysed using SPSS software version 23.0. Qualitative variables were expressed as numbers and frequencies.

## RESULTS

### Pharmacological characteristics of drug-drug interactions

Of the 67 prescriptions with DDIs, 124 interactions were identified, approximately 1.8 interactions per prescription. Pharmacodynamic interactions accounted for 74.2%, pharmacokinetic interactions for 25.8% and physicochemical interactions for none. Combinations to be taken into account (52.5%) represented the most common level of constraint. Among pharmacodynamic DDIs, synergy (89.1%) was the mechanism most frequently encountered, while complexation (59.4%) was predominant in pharmacokinetic interactions.

### Unadvisable combinations

Unadvisable combinations were more common among cardiologists (42.9%) and general practitioners (42.9%). The combination of NSAIDs and NSAIDs was the most common, accounting for 42.9% of cases of inadvisable combinations.

### Drug interactions identified

The majority of drug interactions were not detected by pharmacy staff (78.2%), and personal knowledge (85.2%) was the means of detection most frequently used by staff. The rate of detected interactions was therefore 21.8% in this series.

Pharmacokinetic DDIs were the most frequently detected by pharmacy staff (13.7% of all DDIs). According to the level of constraint, combinations requiring precautionary use (13.7%) were the most frequently detected.

When DDIs were detected, various attitudes were adopted. The majority of dispensers (55.5%) advised the patient on how to use the drugs, and this only happened when a combination requiring precautionary use was involved. As for the combinations to be taken into account, there was no evidence of any attitude on the part of dispensers. The only non recommended drug interaction (3.7%) detected by dispensing staff led to the patient being referred back to the prescriber.

## DISCUSSION

Drug interactions of a pharmacodynamic nature were in the majority with a rate of 74.2%, followed by DDIs of a pharmacokinetic nature with a rate of 25.8%. These results are similar to those of Nguele et al in 2021 on psychotropic drug prescriptions in Cameroon, who found a rate of pharmacodynamic interactions of 72.9% [7]. Pharmacodynamic interactions are more common because they involve drugs with common, complementary or antagonistic pharmacodynamic properties or adverse effects on the same physiological system.

The association to be taken into account (52.5%) represented the level of constraint frequently found. These results corroborate those of Barré et al in 2005, who obtained virtually identical results with 55% of associations to be taken into account in an elderly population hospitalised in a general surgery department in France [8]. This may be explained by the fact that the majority of these interactions were pharmacodynamic and synergistic in nature, which more often corresponds to an addition of adverse effects that the prescriber can monitor during patient follow-up.

The rate of drug interactions detected by pharmacy staff was low (21.8%) and the most common method of detection was personal knowledge (85.2%). This low rate could be explained firstly by the profile of dispensers in our pharmacies, who are not always staff trained for these purposes. Secondly, there are a number of limitations to the detection methods used, which may account for this poor result: variable reliability of memory, tediousness of searching in physical databases facing the crowd of patients.

The associations to be taken into account did not show any attitude adopted by the dispensers. This could be explained by the fact that the risk of drug interaction exists for the combinations to be taken into account, but it is low because it most often corresponds to the addition of side effects. As a result, no practical recommendations can be made, given the sheer number of possible side-effects. Ideally, it is up to the physician to assess the appropriateness of the combination.

Only one out of seven not recommended drug interactions (3.7% of all DDIs) was detected by pharmacy staff and the drug was not dispensed; instead, the patient was referred back to the prescriber. The drug in question was a combination of a potassium-sparing diuretic (spironolactone) and a potassium derivative, with a risk of potentially lethal hyperkalaemia, especially in patients with renal failure. This demonstrates the risk of not looking for DDIs and/or looking for it ineffectively, and above all the importance of good management of DDIs.

## CONCLUSION

The general objective of this study was to describe the management of drug-drug interactions in medical prescriptions received in pharmacies in the city of Yaoundé (Efoulan health district). At the end of the study, it was found that DDIs were mainly pharmacodynamic by synergism, requiring special attention. The detection rate of these possible incidents was low, due to the insufficiently effective techniques used. For the majority, DDIs resulted in advice being given at the same time as the drugs was dispensed. Moreover, the combinations that were not recommended were mainly prescribed by cardiologists and general practitioners. In one case, the patient was referred back to the prescriber because of the lethal risk of the DDI detected.

## Data Availability

Toutes les donnees produites dans la presente etude sont disponibles sur demande raisonnable aupres des auteurs.

## COMPETITING INTEREST

none

---

**Table I:** Pharmaceutical characteristics of drug interactions.

| Variables | Number | Frequency |
| --- | --- | --- |
| **Type of drug interaction** | **N=124** | **%** |
| Pharmacodynamics | 92 | 74.2 |
| Pharmacokinetics | 32 | 25.8 |
| **Level of constraint** | **N=124** | **%** |
| Take into account | 65 | 52.5 |
| Use with caution | 52 | 41.9 |
| Not recommended | 7 | 5.6 |
| **Pharmacodynamic mechanism of action** | **n=92** | **%** |
| Synergism | 82 | 89.1 |
| Potentiation | 7 | 7.6 |
| Antagonism | 3 | 3.3 |
| **Pharmacokinetic mechanism of action** | **n=32** | **%** |
| Complexation | 19 | 59.4 |
| Modification of gastric pH | 5 | 15.6 |
| Renal elimination | 4 | 12.5 |
| Slowing of intestinal peristalsis | 2 | 6.3 |
| Enzyme inhibition | 1 | 3.1 |
| Enzyme induction | 1 | 3.1 |

**Table II:**
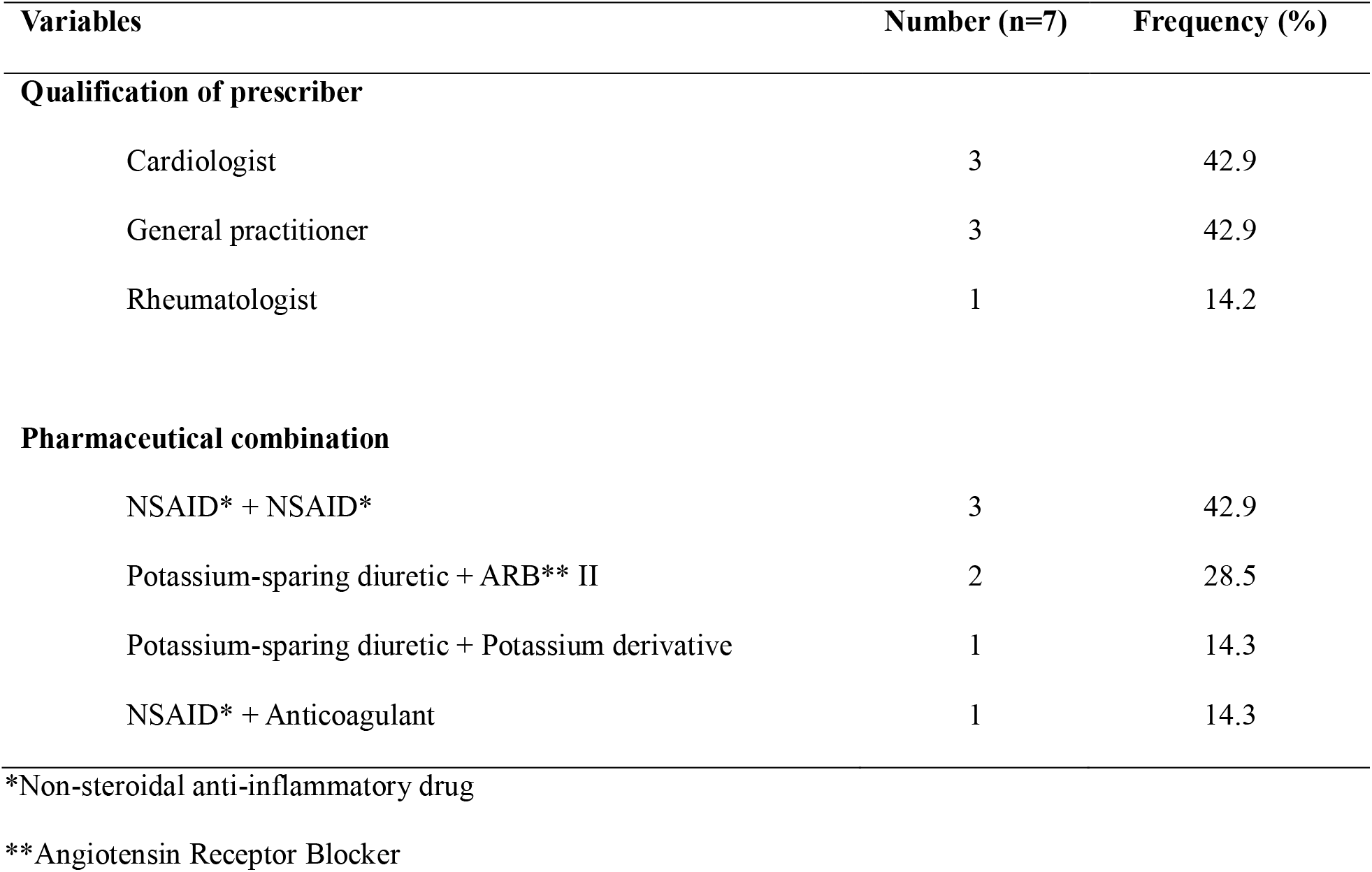
Prescribers and pharmaceutical associations of inadvisable combinations.

**Table III:** Assessment and effective means of detecting.

| Variables | Number | Frequency |
| --- | --- | --- |
| **Drug interaction** | **N=124** | **%** |
| Not detected | 97 | 78.2 |
| Detected | 27 | 21.8 |
| **Effective means of detection** | **n=27** | **%** |
| Personal knowledge | 23 | 85.2 |
| Physical documents | 4 | 14.8 |

**Table IV:** Type and level of constraint according to detection of drug interactions.

| Variables | Interaction detected ? |  |
| --- | --- | --- |
|  | Yes | No |
| **Type of interaction** |  |  |
| Pharmacokinetics | 17 (13.7) | 15 (12.1) |
| Pharmacodynamics | 10 (8.1) | 82 (66.1) |
| **Level of constraint** |  |  |
| Use with caution | 17 (13.7) | 35 (22.2) |
| Take into account | 9 (7.3) | 56 (45.2) |
| Nor recommended | 1 (0.8) | 6 (4.8) |

**Table V:** Attitudes adopted by dispensers according to the level of constraint.

| Attitude adopted | Risk level |  |  |
| --- | --- | --- | --- |
|  | Precaution for use | Take into account | Combination not recommended |
| Advise patient on instructions for use | 15 (55.5) | 0 (0) | 0 (0) |
| No action taken | 2 (7.4) | 9 (33.4) | 0 (0) |
| Refer patient to prescriber | 0 (0) | 0 (0) | 1 (3,7) |

